# THE ROLE OF POINT-OF-CARE ULTRASONOGRAPHY IN THE INITIAL CHARACTERIZATION OF COVID-19 PATIENTS: RESULTS FROM A PROSPECTIVE MULTICENTRIC STUDY

**DOI:** 10.1101/2020.05.15.20103333

**Authors:** Yale Tung-Chen, Rafael Llamas-Fuentes, Pablo Rodríguez-Fuertes

## Abstract

**INTRODUCTION:** Coronavirus Disease 2019 (COVID-19) is a highly contagious illness caused by the Severe acute respiratory syndrome coronavirus 2 (SARS-CoV-2). There is growing evidence regarding the imaging findings of COVID-19, in Chest X-ray and CT scan, however their availability in this pandemic outbreak might be compromised. At this moment, the role of Point-of-care ultrasonography (POCUS) has yet to be explored.

**OBJECTIVES:** The main purpose of this study is to describe the POCUS findings of the disease in COVID-19 patients admitted to the emergency department (ED). Determining the correlation of these parameters with vital signs, laboratory results and chest X-ray, as well as, therapeutic decisions and prognosis.

**METHODS:** Prospective study carried out in the emergency department (ED) of two academic hospitals. High suspicion or confirmed COVID-19 patients were subjected to the ultrasonographic measurement of the inferior vena cava (IVC), focused cardiac ultrasound (FOCUS), and Lung Ultrasonography (Lung POCUS).

**RESULTS:** Between March and April 2020, ninety-six patients were enrolled. The mean age was 68.2 years (SD 17.5). The most common finding in Lung POCUS was an irregular pleural line (63.2%) followed by bilateral confluent (55.2%) and isolated B-lines (53.1%), which was associated with a positive RT-PCR (OR 4.729, 95% CI: 1.989–11.246; p<0.001), and correlated with IL-6 levels (rho = 0.622; p = 0.002). The IVC moderately correlated with levels of pO2, expiratory (rho = –0.539; p = 0.014) and inspiratory (rho = –0.527; p = 0.017), with troponin I (rho = 0.509; p=0.03). After POCUS exam, almost 20% of the patients had an associated condition that required a change in the treatment or management.

**CONCLUSION:** In this pandemic era, as the shortage of resources constitutes an undeniable public health threat, POCUS presents the potential to impact in diagnosis, management and prognosis of our confirmed or suspected COVID-19 patients.

## INTRODUCTION

Coronavirus Disease 2019 (COVID-19) is a highly contagious illness caused by the Severe acute respiratory syndrome coronavirus 2 (SARS-CoV-2). The 11^th^ of March of 2020, the World Health Organization declared a pandemic caused by a novel coronavirus, named Severe Acute Respiratory Syndrome Coronavirus 2 (SARS-CoV-2), with the spread to more than 180 countries (1), 2.274.800 cases confirmed and 156.140 deaths caused (2).

In this emergency, is critical the ability to quickly characterize a confirmed or suspected case, moreover as almost any emergency department will struggle to keep up with the increasing number of patients and the shortage of health resources.

The main diagnostic method is the reverse transcription polymerase chain reaction (RT-PCR) of the nucleic acid of SARS-CoV-2 in nasopharyngeal swabs (3). However, it has many limitations such as the low sensitivity or the technical difficulties to perform it (4).

There have been different studies suggesting that CT abnormalities had a highly sensitivity for diagnosis of COVID-19 patients, and should be considered as a screening tool (4). Moreover, different clinical, laboratory and imaging parameters have been associated with prognosis (5) and to guide therapy (6).

However, since these diagnostic, laboratory and therapy resources may not be ubiquitously available, we need alternative modalities to quicker characterize our patient.

Point-of-care ultrasonography (POCUS) is ubiquitous, is quickly completed following simple and easy to apply protocols (5), therefore it can be performed in mild or even unstable patients, in different settings. The presence of subpleural consolidations, thickened pleural lines and B-lines are highly specific for interstitial syndrome and in these cases suggest the presence of COVID-19 pneumonia (7–8). The role and impact of this technique in this pandemic has not been explored yet.

## PATIENTS AND METHODS

Prospective study carried out in the emergency department (ED) of two academic hospitals. The study was conducted in accordance with the Declaration of Helsinki, and was approved by the Research Ethics Committee of each University Hospital involved. Informed consent was obtained from each enrolled patient.

### Patient selection

Patients admitted to the ED with the clinical suspicion of COVID-19 (temperature above 37.2ºC or acute respiratory symptoms or gastrointestinal symptoms or fatigue) requiring X-ray for evaluation. We excluded patients <18 years or who refused to participate. A convenience sample of patients who met these inclusion criteria were consecutively enrolled and prospectively studied.

Subjects were followed during the following week, either during hospitalization or after hospital discharge, which occurred first.

### Initial patient assessment

Initial evaluation of the patients included recording medical history: demographic data, comorbidities, medications; symptoms; physical exam: temperature, blood pressure, heart rate, respiratory rate and oxygen saturation; Chest X-ray and laboratory tests: hemogram, basic metabolic panel (glucose, electrolytes, kidney function, liver enzymes, etc.), Lactate dehydrogenase (LDH), Ferritin, Interleukin-6 (IL-6), C-Reactive Protein (CRP), Procalcitonin, blood gasses (lactate and pH) and coagulation (D-dimer, INR, PTT, Fibrinogen).

### Ultrasound data collection

Two emergency physicians with long-standing experience in POCUS (experienced sonologists on the basis of the American College of Emergency Physicians ultrasonographic guidelines and more than 10 ultrasound exams performed per week, 5 years of experience in performing and interpreting POCUS (9)) performed all ultrasound exams. Therefore, an opportunity sampling method was implemented for patient selection.

Participants were subjected to ultrasonographic measurement of the inferior vena cava (IVC) and a focused cardiac ultrasound (FOCUS). A Lung Ultrasonography (LUS) was performed following a 12-zone protocol (10). Each intercostal space of upper and lower parts of the anterior, lateral, and posterior regions of the left and right chest wall was carefully examined, and findings (pleural effusion, confluent and isolated B-lines, irregular pleural line, small and lobar consolidations) were recorded (7).

The examinations were performed using a GE LOGIQ-e ultrasound system fitted with a phased and curvilinear array transducer (1.5–4.5 MHz) (General Electrics Healthcare, Madrid, Spain) as a cart-based device and a Butterfly IQ, as a hand-held device.

The physicians were blinded to the patient past medical history, vital signs, symptoms or laboratory measurements.

### Outcome measures and definitions

The main purpose of this study is to describe and characterize the POCUS findings of the disease in COVID-19 patients admitted to the emergency department (ED). The primary outcome was to determine the impact of POCUS parameters to predict the prognosis of patients with high suspicion or confirmed COVID-19. The secondary outcome was to correlate these parameters with the physical exam, laboratory markers and chest X-ray.

We defined a confirmed case any patient with clinical symptoms and positive RTPCR, and high suspicion case to any patient with negative RT-PCR but compatible clinical symptoms and typical X-ray, CT scan or Lung POCUS.

### Statistical analysis

Baseline characteristics are presented as mean and standard deviation (SD) for continuous variables and count and proportions for categorical variables. For group comparisons, we used t-test for continuous variables and the Chi-square or Fisher exact test for categorical one. The correlations between continuous variables were tested using Spearman’s rho test for categorical variables. Mean values were reported along with 95 % confidence intervals. Statistical significance was set at p value < 0.05.

Statistical analyses were conducted with IBM SPSS software v20.0 (SPSS Inc., Chicago, IL, USA).

## RESULTS

Ninety-six patients were enrolled between March and April 2020 (summarized in Table 1). The mean age was 68.2 years (SD 17.5). Fifty patients (52,1%) were female. Nearly half of the patients were hypertensive (49%, 47 patients), being most of them on Angiotensin-converting-enzyme inhibitors (ACEi) or Angiotensin receptor blockers (ARB) therapy. The most common presenting symptom was dyspnea (67.7%) and fever (65.6%), and the mean onset symptoms were 6 days (SD 5.0). The patients were normotensive and had low oxygen saturation (91.8%, SD 6.1), with a respiratory rate of 15 rpm (SD 4.2), needing supplemental oxygen (69.7%). The mean lymphocyte count was 1.34 x10^9 (SD 1.8), C-reactive protein (CRP) of 106.5 (96.8%) and Lactate dehydrogenase of 304 U/L (SD 157.1) at admission. The main therapy was hydroxicholoquine (50 patients, 60.4%). At the end of the first week follow-up, 6 patients had died (6.3%) and 17 were discharged to home (17.1%).

**Table 1:**
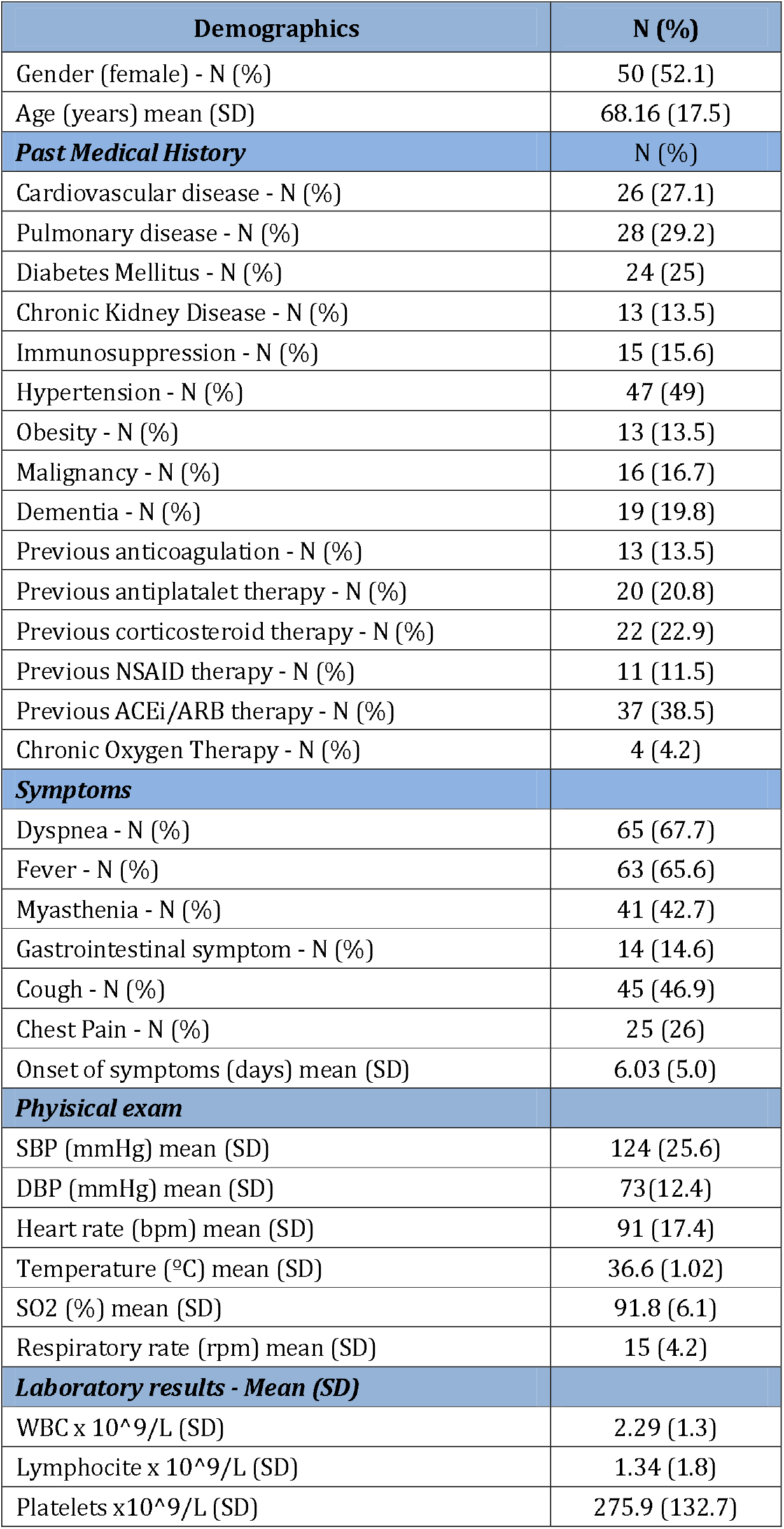
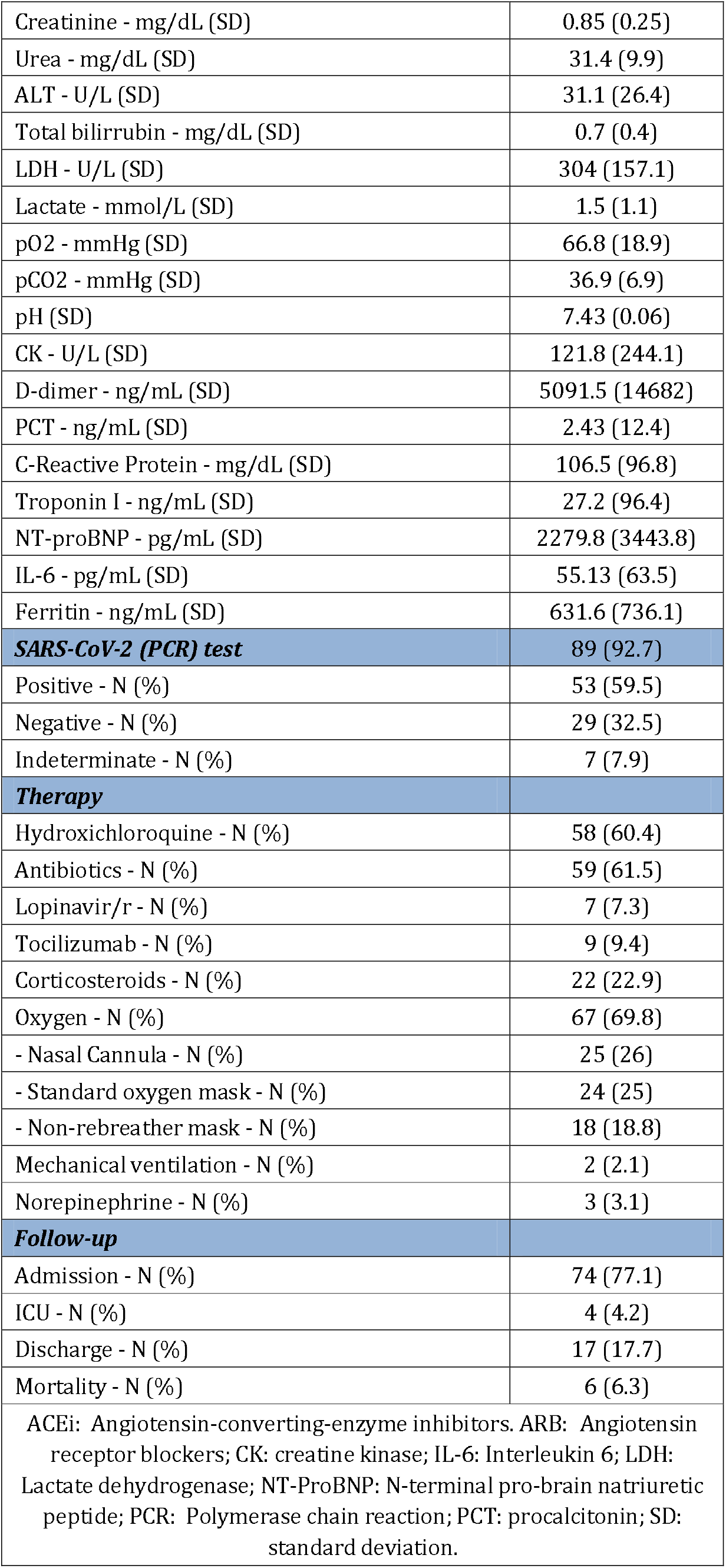
**Demographics and clinical characteristics of patients included (N=96).**

### - Imaging modalities: Chest X-ray and Ultrasound studies

All included patients went through a Point-of-Care ultrasonography (POCUS) study and almost all of them had a chest X-ray (see Table 2).

**Table 2:** **Imaging modalities (Chest X-ray and Point-of-Care Ultrasound) findings of patients included (N=96).**

| Imaging modalities | N (%) |
| --- | --- |
| ***Chest X-ray*** | 95 (99) |
| Normal - (%) | 27 (28.4) |
| Ground-Glass Opacity (GGO) - (%) | 37 (38.9) |
| Interstitial Pattern - (%) | 53 (56.4) |
| Unilobar - (%) | 11 (11.6) |
| Multilobar - (%) | 7 (7.4) |
| Bilateral - (%) | 50 (52.7) |
| ***Point-of-care Ultrasonography (POCUS) results*** | 96 (100) |
| Normal - N (%) | 4 (4.2) |
| Left pleural effusion - N (%) | 15 (15.6) |
| Right pleural effusion - N (%) | 12 (12.5) |
| Bilateral isolated B-lines - N (%) | 51 (53.1) |
| Isolated B-lines - # affected areas (SD) | 2.5 (2.2) |
| Bilateral Confluent B-lines - N (%) | 53 (55.2) |
| Confluent B-lines - # affected areas (SD) | 3.3 (3.2) |
| Bilateral Irregular Pleural Line - N (%) | 61 (63.2) |
| Irregular pleural line - # affected areas (SD) | 3.5 (2.6) |
| Bilateral Small Consolidations - N (%) | 42 (43.8) |
| Small consolidations - # affected areas (SD) | 2.3 (2.4) |
| Pneumonia (lobar consolidation) - N (%) | 1.3 (0.6) |
| Low Ejection Fraction - N (%) | 10 (10.4) |
| Ventricular Dilation - N (%) | 5 (5.2) |
| Atrial dilation - N (%) | 20 (20.8) |
| Ventricular hypertrophy - N (%) | 21 (21.9) |
| Right overload - N (%) | 12 (12.5) |
| Valvulopathy - N (%) | 36 (37.5) |
| Pericardial effusion - N (%) | 16 (16.7) |
| IVC max (mm) (SD) | 14.1 (6.4) |
| IVC min (mm) (SD) | 6.8 (6.2) |
| ***Change/Amendment to diagnosis - N (%) after POCUS*** | 18 (18.8) |
| - Acute pericarditis - N (%) | 4 (4.2) |
| - Decompensated heart failure - N (%) | 4 (4.2) |
| - Bacterial sobreinfection - N (%) | 7 (7.3) |
| - PE/DVT - N (%) | 3 (3.1) |
| DVT: deep vein thrombosis; GGO: Ground-Glass Opacity; IVC: inferior vena cava; PE: pulmonary embolism; POCUS: point-of-care ultrasonography; SD: standard deviation. |  |

The most frequent pattern in chest X-ray was an interstitial pattern (56.4%), and more than a third had ground-glass opacities (GGO). Almost 30% of them had a normal chest X-ray.

Regarding the Lung POCUS, the most common finding was an irregular pleural line(63.2%) followed by bilateral confluent (55.2%) and isolated B-lines (53.1%). 4 patients had a completely normal lung ultrasound. 22 patients (23%) had pleural effusion. The expiratory and inspiratory average diameters of the inferior vena cava (IVC) were 14.1 (6.4) and 6.8 (6.2) mm, respectively. The focused cardiac ultrasound (FOCUS) revealed a low ejection fraction in 10 patients.

After POCUS exam, almost 20% of the patients had an associated condition that required a change in the treatment or management.

### - Correlation of POCUS and RT-PCR of SARS-CoV-2

The presence of bilateral confluent B-lines was associated with a positive RT-PCR (Odds Ratio – OR: 4.729, 95% Confidence Interval – CI: 1.989–11.246, *p*□< 0.001), with a sensitivity (S) of 71.7% and a specificity (Sp) of 65.1%, positive predictive value (PPV) of 61.5% and negative (NPV) of 71.7%. Whereas for chest X-ray had a S of 62.2%, Sp of 71.4%, PPV 88.5% and NPV 34.9% (OR: 4.107, 95% CI: 1.427–11.818, *p*□= 0.006).

The presence of isolated B-lines was associated with a positive RT-PCR (OR: 3.172, 95% CI: 1.145–8.792, *p*□= 0.023), with a S of 86.8% and a Sp of 32.6%, positive PPV of 61.3% and NPV of 66.7%. When considering positive and indeterminate RTPCR it was associated with a S of 81.7%, Sp of 42.9%, PPV 89.3% and NPV 28.5% (OR: 3.350, 95% CI: 1.012-11.094, *p*□= 0.04).

### - Correlation of POCUS and Chest X-ray

The presence of confluent B-lines was associated with pathological findings in the chest X-ray with a S 83.8, Sp 66.7, PPV 89.9 and NPV 53.8 (OR: 10.333, 95% CI: 3.447–30.976, *p* = 0.023), with a S of 86.8% and a Sp of 32.6%, positive PPV of 61.3% and NPV of 66.7%. When considering positive and indeterminate RTPCR it was associated with a S of 81.7%, Sp of 42.9%, PPV 89.3% and NPV 28.5% (OR: 3.350, 95% CI: 1.012–11.094, *p*□<0.001). The presence of consolidations (small and lobar) was associated with a pathological X-ray, with a S 73, Sp 61.9, PPV 87.1 and NPV 39.4 (OR: 4.388, 95% CI: 1.583–12.158, *p*□= 0.003).

Bilateral confluent B-lines was associated with the finding of an interstitial pattern with a S 75.5%, Sp 70,7%, PPV 76.9% and NPV 69% (OR: 2.392, 95% CI: 1.012–5.654, *p*□< 0.001), and ground-glass opacities (GGO) with a S 32.4%, Sp 53.4%, NPV 72.1% and PPV 48.1% (OR: 2.392, 95% CI: 1.012–5.654, *p*□= 0.045).

However, we did not find any significant association with isolated B-lines and interstitial pattern (p=0.156), GGO (0.928) or any pathologic X-ray findings (p=0.831).

### - Correlation of POCUS and Laboratory Parameters

The number of affected areas on Lung POCUS was moderately to strongly correlated to interleukin-6 (IL-6) levels: confluent B-lines (rho = 0.622; p = 0.002), irregular pleural line (rho = 0.509; p = 0.013); as well as a high inspiratory IVC (rho = 0.550; p=0.007). A pathologic Chest X-ray showed a lower correlation (rho = 0.442; p=0.035). Other laboratory and inflammatory markers showed a good correlation with IL-6: CRP (rho = 0.604; p=0.002), Procalcitonin (rho = 0.504; p=0.024), Ferritin (rho = 0.579; p=0.005), AST (rho = 0.635; p=0.001) and LDH (rho 0.695; p<0.001). The highest correlation was found with the respiratory rate (rho = 0.789; p<0.001) and NT-proBNP (0.990; p=0.001). These patients were more prone to receive an anti-IL-6 therapy (rho = 0.612; p=0.002).

The IVC moderately correlated with levels of pO2, expiratory (rho = –0.539; p=0.014) and inspiratory (rho = –0.527; p=0.017), with troponin I (rho = 0.509; p=0.03).

Patients with more comorbid diseases were more prone to have apical lungs involvement: hypertension (OR: 3.040, 95% CI: 1.055–8.762, p=0.034), cardiomyopathy (OR: 2.917, 95% CI: 1.152–7.386, p=0.021) and dementia (OR: 4.286, 95% CI: 1.492–12.310, p=0.005). Although this had a weak correlation with poor outcome (rho = 0.217; p=0.034), such as mortality or need of mechanical ventilation, it was comparable to lymphocyte count (rho = –0.273; p=0.009), creatinine (rho = 0.267; p=0.011) and Procalcitonin (rho = 0.367; p=0.002).

### - Correlation of POCUS and Therapy

Patients with confluent B-lines had higher chances to receive anti-IL-6 therapy (rho = 0.206; p=0.045), and lobar consolidation with hydroxicholoroquine (rho = 0.810; P = 0.001). Remarkably this correlation was much lower when small consolidations (rho = 252, p=0.013), confluent (rho= 0.262; p=0.01) or isolated (rho = 0.279, p=0.006) B-lines were present.

## DISCUSSION

Safety and quality are vital components in ED patient’s management. Many hospitals are struggling to reduce ED overcrowding and increase patient safety through multimodal interventions on patient flow in the ED, especially with laboratory and diagnostic imaging departments (11).

There is growing literature regarding the prognostic factors (5), diagnosis (3–4) and therapeutic challenges (6) in COVID-19 patients.

### - Diagnosis

The positivity rate of RT-PCR has been quantified as 63% in nasal swab and 32% in pharyngeal swab (12), similar to our results, we found a positive rate of only 59.5%. Due to its limitations, diagnostic imaging plays a key role in the management of these patients.

A study of 1049 patients undergoing chest CT scan and RT-PCR testing determined that CT abnormalities had a highly sensitivity for diagnosis of COVID-19 patients (4), suggesting that CT scan should be considered as a screening tool, especially in epidemic areas with high pre-test probability. However, the use of CT scan in the ED has many limitations, such as the radiation exposure, especially for mild illness, the low availability and the contraindication of its use in unstable patients.

Therefore in many centers CT scans have been replaced for chest X-ray. However, as we have seen, chest X-ray has shown to have a very low NPV (34.9%). In a study of patients undergoing an initial screening for COVID-19, they found a sensitivity of 25% and a specificity of 90% (13).

In our study we found that 27 patients with normal chest X-ray, 23 (85.1%) had a pathological POCUS finding. A previous study found that a normal chest X-ray was present in 31% of COVID-19 RT-PCR positive patients (14), which is similar to our results (28.4%). We hypothesize that this is due to the low accuracy of X-ray for detecting interstitial abnormalities (5), represented in our study as isolated B-lines on Lung POCUS, and becoming apparent on X-ray as the disease progresses, with the appearance of confluent B-lines and other findings.

### - Therapy

By adding POCUS to our protocol, we could safely exclude the probability of other synchronous or comorbid diseases, such as deep vein thrombosis, pericardial effusion, heart failure or lobar pneumonia (highly suggestive of bacterial origin), in our study seen in approximately 1 out of 5 patients (18.8%). These findings should trigger the initiation or adjustment of therapy (i.e. antibiotics, anticoagulants, diuretics or, even, colchicine).

Moreover, we showed that that the number of affected lung areas correlates with inflammatory markers, such as IL-6, which in turned could serve as a guide to start therapy with an anti-IL-6 therapy (i.e. tocilizumab). Remarkably, but probably expected, this marker was associated with a higher respiratory rate, acute phase reactants (CRP, Procalcitonin, Ferritin) and LDH, which according to previous studies are also prognostic markers (5). Although we did not see a correlation with ICU admission, therapeutic or invasive procedures or death, this could be due to the short follow-up (one week).

### - Follow-up and prognosis

There has been proposed that changes in the proportion of CT scan GGO lesion, crazy paving pattern and consolidation varies with time and disease progression (15–16), which could be a marker of the stage of the disease. As this disease tends to rapid progression, CT scan may not be available or the patient condition does not allow to perform it (14). As previously reported in COVID-19 era, there is a correlation of Lung POCUS findings to those of the CT scan (17–18), therefore follow-up could be more easily replaced with POCUS as it would be more accessible, and should be explored in future studies.

The presence of apical lung involvement on POCUS correlates in our study with different comorbid diseases (hypertension, cardiomyopathy, dementia) comparable to that yield by different laboratory markers (creatinine, lymphocyte count or procalcitonin), and as expected, in the prognosis (5).

The IVC, as a marker of fluid status, moderately correlated with levels of pO2 and troponin I, which could represent a situation of hemodynamic congestion and worse oxygenation. Therefore, we believe that integrating the IVC in our current practice is appropriate as it addresses more physiologically the assessment of the volume status.

In our study we found a higher prevalence of pleural effusion (23%) than previously reported (14, 16), this could be due to the accuracy of the technique, compared to CT scan or chest X-ray (19), therefore its mere presence should not be considered as a prognostic factor.

### - Strengths

To our knowledge, this is the first study evaluating the potential impact of POCUS in COVID-19 patients, with diagnostic, prognostic and therapeutic implications.

We want to share our study findings, given the urgent need for different strategies in order to better manage COVID-19 patients, and diminish the SARS-CoV-2 spread and its prognosis in the current pandemic context. As the shortage of resources constitutes an undeniable public health threat, we consider POCUS to be a potential solution, and recommend that it should be performed as a first-line imaging test for COVID-19 patients.

### - Limitations

There are several limitations to consider. The main limitation is that Lung POCUS findings overlap with those from other pneumonia etiologies. However, in epidemic areas, positive Lung POCUS features, even in negative RT-PCR or chest X-ray can still be highly suggestive of COVID-19 infection, which could preclude that the sensitivity and specificity reported of Lung POCUS might be higher, and therefore more studies should be carried, comparing with other techniques (i.e. CT scan). Many COVID-19 patients in our ED, with negative RT-PCR or chest X-ray, do not always get a chest CT performed, and therefore there is a chance of misdiagnosis. Minimized, as the patients were followed-up, by reviewing their electronic history, and any complication was recorded.

Another limitation is that selection bias might have occurred. Two experts sonographers performed all ultrasound scans on a consecutive sample selected based on their availability (during their working hours), which limits the generalizability of our results. The impact of this limitation is minimized by the variable schedule and changing shifts, unpredictable a priori (in continuous care). Additionally, false negative ultrasound might be found in the initial stage of the disease, before lung involvement.

Thus, the results from this study open an opportunity to further investigate the use of ultrasound in different settings and clinical scenarios.

## CONCLUSIONS

In this pandemic era, as the shortage of resources constitutes an undeniable public health threat, Point-of-Care Ultrasonography presents the potential to impact in diagnosis, management and prognosis of our confirmed or suspected COVID-19 patients.

## Data Availability

Data accesible on request to

## Acknowledgments

- The authors have declared no conflicts of interest.

## REFERENCES

1. World Health Organization. Rolling updates on coronavirus (COVID-19). WHO characterizes COVID-19 as a pandemic. (https://www.who.int/emergencies/diseases/novel-coronavirus-2019/events-as-they-happen)

2. Johns Hopkins Coronavirus Resource Center. Available at: https://coronavirus.jhu.edu/map.html. (Accessed: 18th April 2020)

3. Guidance for Corona Virus Disease 2019: Prevention, Control, Diagnosis and Management National Health Commission (NHC) of the PRC, General Office; National Administration of Traditional Chinese Medicine of the PRC, General Office. Available at: http://www.pmph.com/patients.

4. Ai T, Yang Z, Hou H, Zhan C, Chen C, Lv W, et al. Correlation of Chest CT and RT-PCR Testing in Coronavirus Disease 2019 (COVID-19) in China: A Report of 1014 Cases. Radiology. 2020 Feb 26;200642.

5. Guan W, Ni Z, Hu Y, Liang W, Ou C, He J, et al. Clinical Characteristics of Coronavirus Disease 2019 in China. N Engl J Med. 2020 Feb 28;NEJMoa2002032.

6. Sanders JM, Monogue ML, Jodlowski TZ, Cutrell JB. Pharmacologic Treatments for Coronavirus Disease 2019 (COVID-19): A Review. JAMA [Internet]. 2020 Apr 13 [cited 2020 Apr 14]; Available from: https://jamanetwork.com/journals/jama/fullarticle/2764727

7. Volpicelli G, Lamorte A, Villén T. WHAT’S NEW IN LUNG ULTRASOUND DURING THE COVID-19 PANDEMIC. Intensive Care Med. 2020;16.

8. Huang C, Wang Y, Li X, Ren L, Zhao J, Hu Y, et al. Clinical features of patients infected with 2019 novel coronavirus in Wuhan, China. The Lancet. 2020 Feb;395(10223):497–506.

9. Ultrasound Guidelines: Emergency, Point-of-Care and Clinical Ultrasound Guidelines in Medicine. Ann Emerg Med. 2017 May;69(5):e27–54.

10. Soummer A, Perbet S, Brisson H, Arbelot C, Constantin J-M, Lu Q, et al. Ultrasound assessment of lung aeration loss during a successful weaning trial predicts postextubation distress*: Crit Care Med. 2012 Jul;40(7):2064–72.

11. Shaw JLV. Practical challenges related to point of care testing. Pract Lab Med. 2016 Apr;4:22–9.

12. Wang W, Xu Y, Gao R, Lu R, Han K, Wu G, et al. Detection of SARS-CoV-2 in Different Types of Clinical Specimens. JAMA [Internet]. 2020 Mar 11 [cited 2020 Apr 18]; Available from: https://jamanetwork.com/journals/jama/fullarticle/2762997

13. Choi H, Qi X, Yoon SH, Park SJ, Lee KH, Kim JY, et al. Extension of Coronavirus Disease 2019 (COVID-19) on Chest CT and Implications for Chest Radiograph Interpretation. Radiol Cardiothorac Imaging. 2020 Apr 1;2(2):e200107.

14. Wong HYF, Lam HYS, Fong AH-T, Leung ST, Chin TW-Y, Lo CSY, et al. Frequency and Distribution of Chest Radiographic Findings in COVID-19 Positive Patients. Radiology. 2019 Mar 27;201160.

15. Pan F, Ye T, Sun P, Gui S, Liang B, Li L, et al. Time Course of Lung Changes On Chest CT During Recovery From 2019 Novel Coronavirus (COVID-19) Pneumonia. Radiology. 2020 Feb 13;200370.

16. Bernheim A, Mei X, Huang M, Yang Y, Fayad ZA, Zhang N, et al. Chest CT Findings in Coronavirus Disease-19 (COVID-19): Relationship to Duration of Infection. Radiology. 2020 Feb 20;200463.

17. Poggiali E, Dacrema A, Bastoni D, Tinelli V, Demichele E, Mateo Ramos P, et al. Can Lung US Help Critical Care Clinicians in the Early Diagnosis of Novel Coronavirus (COVID-19) Pneumonia? Radiology. 2020 Mar 13;200847.

18. Chinese Critical Care Ultrasound Study Group (CCUSG), Peng Q-Y, Wang X-T, Zhang L-N. Findings of lung ultrasonography of novel corona virus pneumonia during the 2019–2020 epidemic. Intensive Care Med [Internet]. 2020 Mar 12 [cited 2020 Apr 4]; Available from: http://link.springer.com/10.1007/s00134-020-05996-6

19. Volpicelli G, Elbarbary M, Blaivas M, Lichtenstein DA, Mathis G, Kirkpatrick AW, et al. International evidence-based recommendations for point-of-care lung ultrasound. Intensive Care Med. 2012 Apr;38(4):577–91.

